# Prediction of Parkinson’s disease progression from sparse longitudinal trajectories using multiclass likelihood contrast learning

**DOI:** 10.64898/2026.09.07.26362404

**Authors:** Sangam Pangeni, Md Rejuan Haque, Fahad Mostafa

**Author notes:** Corresponding /.

## Abstract

**Background:** Biomedical studies increasingly collect repeated measurements, such as clinical ratings, speech measures, gait summaries, and wearable-sensor features, at irregular subject-specific follow-up times. These data are often sparse and unaligned, making them difficult to use with standard classifiers that require fixed-length input vectors. Parkinson’s disease (PD) provides a motivating example because progression is longitudinal and heterogeneous, but clinical follow-up is rarely observed on a common schedule.

**Objective:** This study develops and evaluates a multiclass likelihood contrast classifier for sparse longitudinal trajectory recovery. The goal is to classify subjects using their observed repeated measurements while avoiding aggressive reduction of trajectories to averages or simple slopes.

**Methods:** The proposed method fits one class-specific mixed-effects model per outcome class and assigns each held-out subject to the class with the largest marginal likelihood score. The main specification uses a quadratic mean trajectory with subject-specific random intercept and random slope terms. Performance is evaluated in a three-class simulation with imbalanced class sizes and in a Parkinson’s motor-UPDRS sparse trajectory recovery experiment. In the Parkinson’s experiment, dense histories are first used to define mathematical trajectory phenotypes; most observations are then hidden, and models are asked to recover the phenotype from the sparse record. Baselines include a linear mixed-likelihood classifier, functional k-nearest neighbors, random forest, and support vector machine.

**Results:** In the Parkinson’s sparse recovery experiment, after approximately 90% of dense observations were hidden, the proposed model achieved accuracy 0.8095, macro-F1 0.8160, MCC 0.7107, and macro-AUC 0.9268. A train–validation–test diagnostic gave similar held-out behavior, with validation macro-F1 0.7980 and test macro-F1 0.8160 for the proposed model.

**Conclusion:** Multiclass likelihood contrasts offer an interpretable and competitive approach for sparse, unaligned longitudinal classification. The Parkinson’s analysis should be read as an algorithmic sparse trajectory recovery study rather than clinical diagnostic validation. Future work should evaluate the method with externally assigned clinical outcomes, multivariate longitudinal biomarkers, prospective cohorts, and ordinal generalized mixed-model likelihoods that better match bounded clinical rating scales.

## 1 Introduction

Longitudinal data play a central role in Parkinson’s disease research because disease progression cannot be adequately characterized by a single clinical assessment. PD is a chronic neurodegenerative disorder marked by substantial heterogeneity in symptom onset, progression rate, and treatment response. Motor manifestations, including tremor, rigidity, bradykinesia, and postural instability, often evolve gradually over time, while non-motor symptoms may follow distinct trajectories across patients. Repeated measurements of clinician-rated outcomes, such as the Movement Disorder Society Unified Parkinson’s Disease Rating Scale (MDS-UPDRS), together with speech assessments, gait characteristics, cognitive measures, and wearable-sensor biomarkers, provide valuable information about disease progression. Capturing these temporal patterns is essential for understanding disease heterogeneity, identifying progression phenotypes, and developing accurate predictive models for patient stratification and prognosis [12, 15, 26, 40]. This longitudinal structure is scientifically valuable because repeated measurements can reveal progression patterns that are invisible at a single visit. At the same time, it creates a practical statistical problem: real patients rarely contribute data on a common schedule. They may enter a study at different stages, miss scheduled visits, leave the study early, or contribute additional assessments at irregular times.

The resulting data set is usually not a rectangular matrix. One subject may have four observations, another may have twenty, and the observation times may not match. Standard supervised learning methods are often built around fixed-length feature vectors, so analysts must first convert the repeated measurements into a subject-level representation. A common strategy is to compute simple summaries such as the mean, standard deviation, minimum, maximum, last value, first-to-last change, or fitted linear slope. These summaries make the data compatible with familiar classifiers such as logistic regression, random forests, support vector machines, and nearest-neighbor methods [6, 9, 25]. The price is that important temporal information may be lost. Two subjects can have similar averages and similar slopes even when one follows a U-shaped trajectory and the other follows a monotone trend. When the scientific signal is curvature, acceleration, relapse, or recovery, fixed summaries may not represent the relevant pattern.

A second common strategy is to impute or interpolate all subjects onto a shared grid. This can be useful, particularly when the underlying process is smooth and the observations are reasonably dense. However, grid construction may introduce artifacts when follow-up is highly sparse, when visit times are informative, or when subjects have very different observation windows. The problem is especially important in biomedical settings where the number of subjects may be modest and where a few visits per subject must carry substantial information. For this reason, methods that can use the repeated-measures table directly remain attractive. They allow analysts to preserve the actual visit times and avoid pretending that every subject was observed in the same way.

Recent machine learning research has proposed many models for irregularly sampled time series. Recurrent neural networks with decay mechanisms and time-aware recurrent models have been used to handle missingness and irregular time gaps [4, 7]. Set-function approaches, latent ordinary differential equation models, and neural controlled differential equations offer another way to represent asynchronous measurements [13, 16, 33]. Graph-based models, time-parameterized convolutions, and transformer-based temporal architectures have also been developed for irregular clinical time series [18, 21, 44]. Recent surveys and newer sequence models continue to expand the range of deep learning tools available for sparse health data [20, 37, 43]. These approaches are promising for large-scale health records and sensor streams. Nevertheless, highly parameterized models may require large sample sizes, extensive hyperparameter tuning, careful calibration, and substantial computational infrastructure. In smaller biomedical studies, interpretable likelihood-based models remain valuable because their assumptions are explicit and their outputs are easier to connect to the scientific question.

Mixed-effects models provide one of the most established statistical frameworks for irregular longitudinal data. They naturally allow unequal numbers of observations per subject, nonaligned time points, and within-subject correlation through random effects [3, 10, 19, 41]. They are widely used for clinical progression modeling, longitudinal regression, joint modeling, and latent class analysis [28, 30, 36]. Their usual role is estimation and inference, but the likelihood from a fitted mixed model can also be used for classification. The likelihood contrast approach follows this logic: fit one longitudinal model for each group, evaluate a new subject under each group-specific model, and assign the subject to the group that gives the most favorable likelihood contrast [17]. The original likelihood contrast work focused on binary classification. The same decision principle extends naturally to multiple classes by fitting one model for each class and comparing all class-specific likelihood scores.

Functional data analysis provides another relevant tradition. Functional methods treat observations over time as noisy measurements of an underlying curve and include tools for sparse functional principal components, functional regression, curve classification, and trajectory distance comparisons [5, 14, 29, 42]. These methods are important because they address trajectory shape more directly than scalar summaries. In this study, we therefore include a functional k-nearest-neighbor baseline. This is a stronger contextual comparator than only using random forests or support vector machines on summary features, because functional kNN attempts to classify subjects from estimated trajectory shape.

Since sparse and irregularly observed longitudinal data are common in clinical studies of Parkinson’s disease and other chronic conditions. Existing classification approaches often fail to fully exploit the temporal structure of these data because they require aligned observations or rely on summary measures. The central challenge addressed in this study is how to accurately classify subjects and recover underlying progression trajectories from limited, unaligned longitudinal observations while maintaining model interpretability. The main contributions of this study are summarized as follows:

- Propose a multiclass likelihood contrast classification framework for sparse and irregularly observed longitudinal data based on class-specific mixed-effects models.
- Develop a trajectory recovery approach that leverages the full longitudinal structure of repeated measurements rather than relying on summary statistics or fixed-length feature representations.
- Introduce a Parkinson’s disease sparse trajectory recovery experiment to evaluate the ability of classification models to recover latent progression phenotypes from limited follow-up observations.
- Compare the proposed method with established statistical and machine-learning approaches, including functional k-nearest neighbors, random forest, and support vector machine classifiers, and demonstrate competitive predictive performance in both simulation and real-data settings.

Therefore, first, we present a direct multiclass extension of likelihood contrast classification for sparse longitudinal trajectories. Second, we evaluate it under subject-level cross-validation against both standard fixed-vector baselines and a functional time-series baseline. Third, we report the Parkinson’s analysis transparently as sparse trajectory recovery, including subject counts, class sizes, observation counts, and masking levels.

## 2 Data Introduction and Preprocessing

The empirical application uses the Parkinson’s telemonitoring data set introduced by Tsanas et al. [40]. The original table contains 5,875 repeated observations from 42 individuals with Parkinson’s disease. Each record includes subject identifier, age, sex, follow-up time, motor-UPDRS, total-UPDRS, and several speech-derived acoustic variables such as jitter, shimmer, noise-to-harmonics ratio, harmonics-to-noise ratio, recurrence period density entropy, detrended fluctuation analysis, and pitch period entropy. In the present analysis, motor-UPDRS is the longitudinal response and test time is the time variable because the goal is to study recovery of motor-severity trajectory phenotypes rather than to build a multivariate speech-based diagnostic model. The motor-UPDRS values range from 5.04 to 39.51, with mean 21.30 and standard deviation 8.13, while follow-up times range from approximately -4.26 to 215.49 days. Subjects contribute a dense record with a mean of about 140 observations per subject, but the recovery experiment keeps only 587 sparse observations, approximately 14 per subject. The dense histories are used to define three trajectory phenotypes with class sizes 10, 10, and 22 subjects. All preprocessing, including standardization of time and response values, is performed within subject-level training folds to avoid leakage into validation or test subjects [12, 17, 40].

## 3 Methodology

This study evaluated the proposed multiclass likelihood contrast framework through both simulation experiments and a Parkinson’s disease sparse trajectory recovery application. We first conducted a controlled simulation study to assess the ability of the method to recover multiclass longitudinal trajectory patterns under sparse and irregular observation schedules. The proposed model was then trained and evaluated using longitudinal motor-UPDRS trajectories from individuals with Parkinson’s disease, where dense histories were used to define trajectory phenotypes and sparse observations were used for classification. The following sections describe the problem formulation, class-specific mixed-effects likelihood model, data preprocessing procedures, validation strategy, comparator methods, and performance evaluation metrics used throughout the simulation and Parkinson’s disease analyses[10, 17, 19, 41] (see in Figure 1).

**Figure 1:**
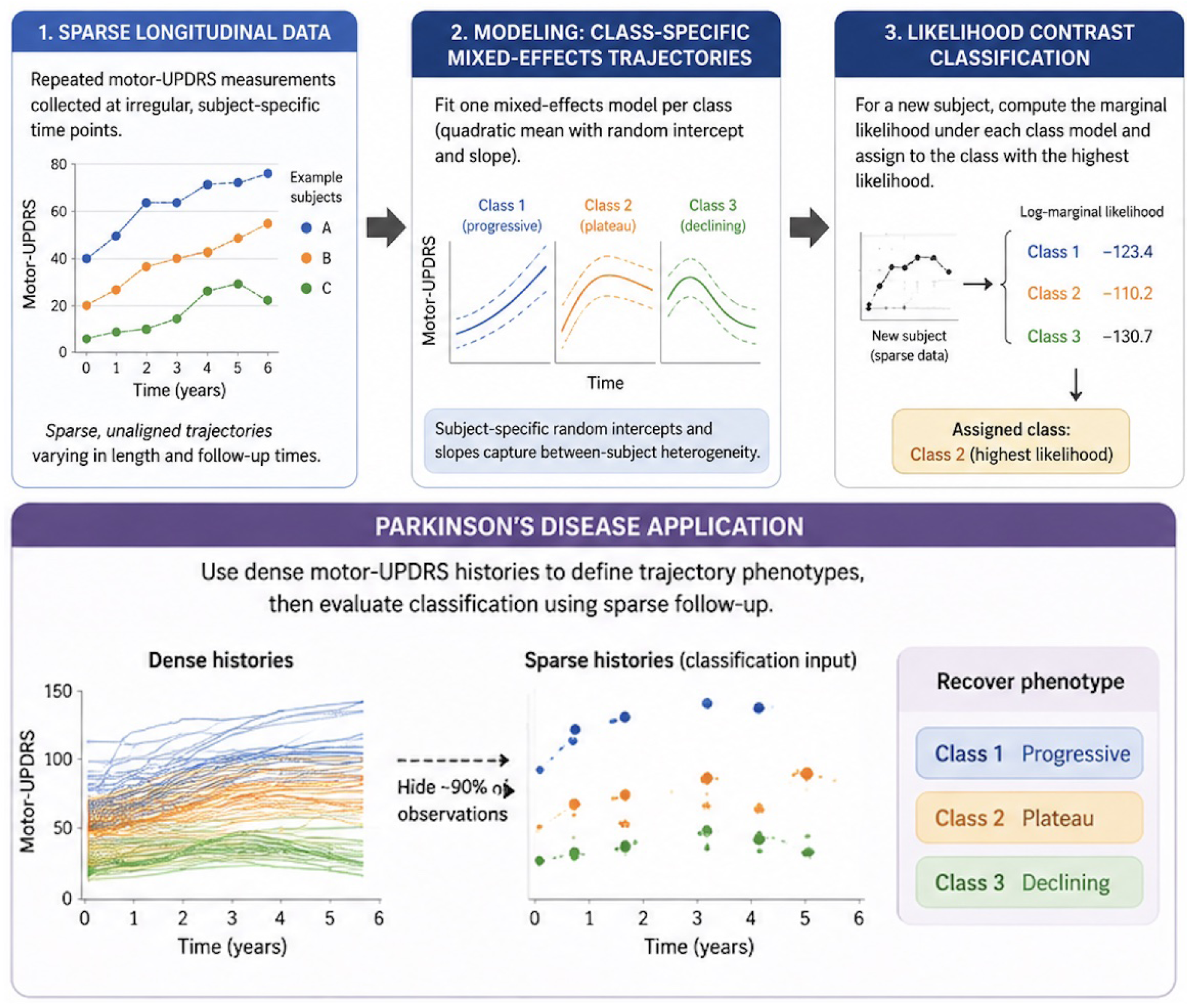
The proposed multiclass likelihood contrast learning framework for sparse longitudinal trajectory recovery in Parkinson’s disease.

### 3.1 Problem setup

Let subject *i* = 1*, . . . , N* have repeated measurements (*t_ij_, y_ij_*) for *j* = 1*, . . . , n_i_*, where *t_ij_*is the observation time and *y_ij_* is the longitudinal response. The number of observations *n_i_* can vary by subject, and the time points do not need to be aligned across subjects. Each subject belongs to one of *K* classes, denoted *C_i_* ∈ {1*, . . . , K*}. In our experiments, *K* = 3. The goal is to predict *C_i_* for a held-out subject using only the subject’s sparse measurements [10, 41].

The key modeling decision is to classify at the subject level rather than at the visit level. Visit-level splitting would place observations from the same subject into both training and testing sets, producing overly optimistic estimates because the test observations would not be independent of the training observations. We therefore split subjects, not visits. All observations from a given subject remain together in either the training fold or the test fold [19, 28].

### 3.2 Class-specific mixed-effects likelihood model

For each class *k*, the proposed classifier fits a class-specific linear mixed-effects model [3, 19, 41]. In the quadratic specification used as the main model, the measurement vector for a subject is represented as

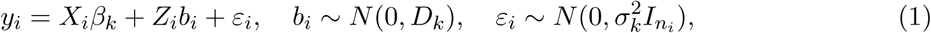

where *X_i_* contains the fixed-effect columns (1*, t, t*^2^), *Z_i_* contains the random-effect columns (1*, t*), *β_k_* is the class-specific mean trajectory parameter, *D_k_* is the class-specific random-effect covariance matrix, and *σ_k_*^2^ is the residual variance. After integrating over the random effects, the marginal distribution for subject *i* under class *k* is

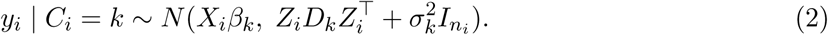

This marginal distribution is what allows the classifier to score a subject with any number of observed visits at any observed times [10, 30].

For prediction, the fitted model for each class gives a log-likelihood score

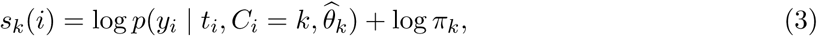

where 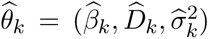 and *π_k_* is the class prior. We use uniform priors in the main analysis because the goal is balanced multiclass performance rather than reproducing the empirical class imbalance [17, 38]. The predicted class is

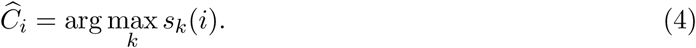

Posterior-like class probabilities are obtained by applying a numerically stable softmax to the class log-scores. These probabilities are used for macro-AUC. Models are fitted by maximum likelihood rather than restricted maximum likelihood so that likelihoods are comparable across class-specific fixed-effect structures [3, 36]. In small folds, a mixed-effects fit can occasionally encounter singular covariance estimates or optimizer failures. The implementation therefore uses conservative numerical safeguards: covariance matrices are symmetrized and shifted to the nearest positive semi-definite form when needed, and a class-specific Gaussian polynomial fallback is used if a mixed-effects fit fails. This safeguard prevents a single failed class fit from producing unusable infinite scores while keeping the decision rule likelihood-based.

#### Algorithm 1

**Multiclass mixed-effects likelihood contrast classification**

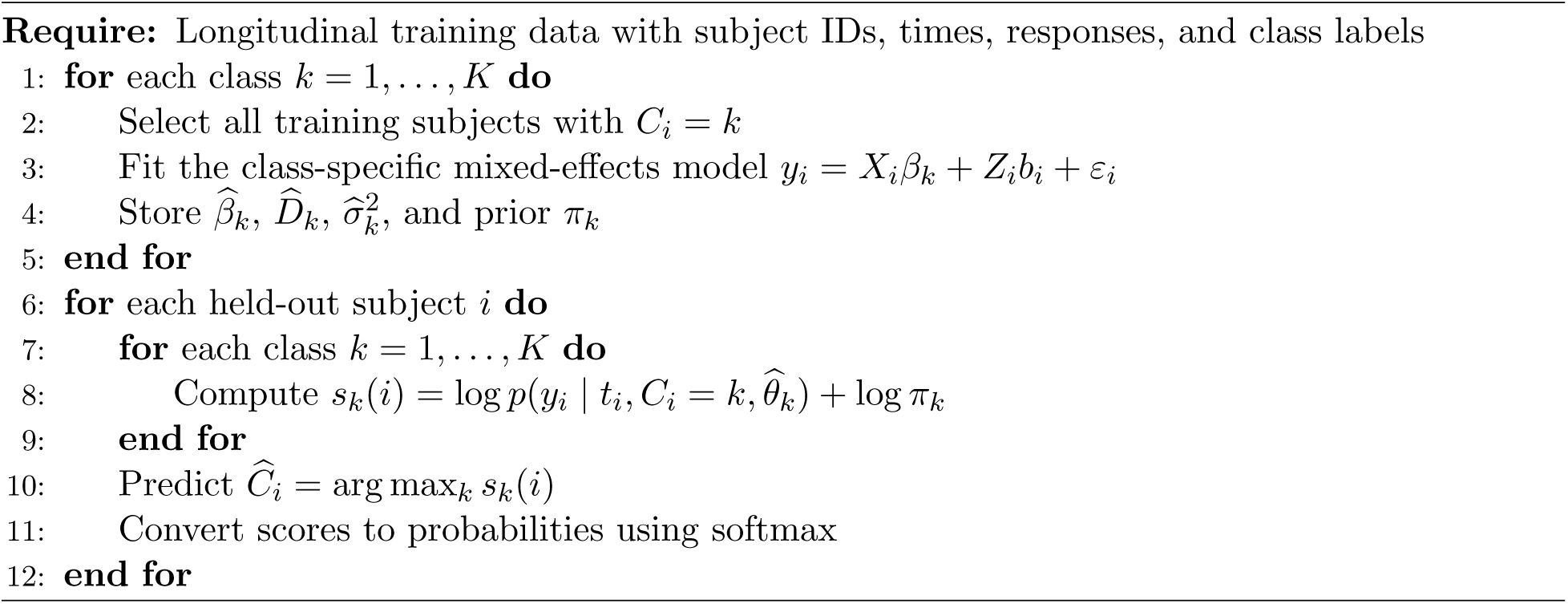

### 3.3 Data preprocessing and validation protocol

All models are evaluated using stratified five-fold cross-validation at the subject level [17, 25]. Stratification is applied to subject labels, and all visits from a subject are kept in the same fold. Before model fitting within each fold, time and response values are standardized using the training fold only [25]. The same training-fold mean and standard deviation are then applied to the corresponding test fold. This prevents leakage of test-fold information into the fitted models and makes the mixed-effects optimization more stable. The mixed-likelihood models receive the repeated-measures data directly. The random forest and support vector machine receive subject-level summaries extracted after fold-wise normalization.

The Parkinson’s sparse recovery experiment uses a two-stage design. First, dense histories are used only to define trajectory phenotypes. Subject-level polynomial descriptors are computed from the dense trajectories and clustered into three groups with *k*-means. Second, sparse inputs are created by retaining only a subset of each subject’s visits. Models are trained and evaluated only on the sparse repeated-measures table [26, 40]. This design deliberately separates phenotype construction from sparse-input classification, but it does not create independent clinical labels. For that reason, the real-data experiment is reported as a recovery experiment rather than a clinical validation study.

### 3.4 Baseline models

We compare the proposed quadratic multiclass MLC model with four baselines. The first baseline is a linear mixed-likelihood classifier with a linear class mean and subject-specific random intercept. This model tests whether the quadratic mean and random slope improve recovery of curved trajectories. The second baseline is functional k-nearest neighbors, motivated by functional data analysis and curve-classification ideas [14, 29, 42]. For each fold, sparse subject trajectories are interpolated onto a grid determined by the training fold, distances are computed between interpolated curves, and the class label is predicted from neighboring training trajectories. This baseline is included because functional methods are specifically designed for curve-like data and are therefore a relevant comparison for irregular longitudinal classification.

The remaining two baselines are random forest and radial-basis support vector machine classifiers [6, 9, 25]. These models are trained on common subject-level summaries: mean, standard deviation, minimum, maximum, and linear slope. These fixed-vector summaries represent a common machine-learning workflow for longitudinal biomedical data. They also illustrate the cost of compressing a sparse trajectory into a small number of scalar descriptors.

### 3.5 Evaluation metrics

We report four metrics: accuracy, macro-F1, MCC, and macro-AUC. Accuracy is the overall proportion of correctly classified subjects. Macro-F1 averages class-specific F1 scores and therefore gives equal weight to each class. MCC uses the full multiclass confusion structure and is informative under class imbalance. Macro-AUC evaluates one-vs-rest probability ranking quality averaged across classes. These metrics are computed from pooled held-out predictions across the five cross-validation folds.

### 3.6 Reproducibility and leakage controls

Several design choices were used to reduce optimistic bias. First, the unit of prediction is always the subject, not the visit. Second, normalization is estimated only from the training fold in each split. Third, Parkinson’s trajectory phenotypes are constructed from dense histories before sparsification, but model training and testing use only sparse inputs. This makes the recovery target explicit and prevents the classifier from seeing the dense record at prediction time [17, 40]. Fourth, all random operations are seeded for reproducibility. The global seed is RANDOM STATE = 42; the outer cross-validation uses this seed, the Parkinson’s inner validation split uses 42 + fold, and the simulation sample-size study uses 42 + 100N + replicate for each total sample size *N* . The main validation strategy is not simple random sampling: it uses stratified subject-level five-fold cross-validation with shuffle=True, so each fold uses approximately 80% of subjects for model fitting and 20% for held-out testing while preserving class proportions as closely as possible [25]. For the additional train–validation–test diagnostic in the Parkinson’s experiment, 20% of each training fold is set aside using stratified subject-level sampling, giving an approximate 64%/16%/20% training/validation/test structure by subject.

No hyperparameters are tuned on the test folds. The reported values are fixed analysis settings selected before evaluation from the modeling design and kept unchanged across experiments, not estimates chosen by a test-set grid search. In both the simulation and Parkinson’s sparse recovery analyses, the proposed MLC uses a quadratic fixed mean (degree = 2), random intercept and random time slope (random effects=intercept slope), uniform class priors, ridge stabilization 10^−4^, and covariance shrinkage 0.0. The linear MLC baseline uses degree = 1, a random intercept, uniform priors, ridge stabilization 10^−4^, and covariance shrinkage 0.0. Functional kNN uses five neighbors, distance weighting, and a ten-point interpolation grid. The random forest uses 160 trees, minimum leaf size 2, and balanced subsampling class weights. The radial-basis SVM uses *C* = 2.0, gamma=scale, one-vs-rest decision scores, and balanced class weights [6, 9, 25].

The simulation study additionally fixes the data-generating settings at a two-year time window, 4–8 visits per subject, Gaussian noise standard deviation 0.45, a 10% visit-level missingness step when feasible, random-intercept standard deviation 0.70, random-slope standard deviation 0.25, and class proportions close to 1:2:3 for *N* = 50, 100, 300, 500, and 1000. The Parkinson’s sparse recovery study fixes the retained fraction at 0.10, uses *k* = 3 trajectory phenotypes from dense-history descriptors, and fits *k*-means with 50 initializations and the same global random seed [22, 31].

### 3.7 Statistical and computational considerations

The computational burden of the proposed approach comes mainly from fitting one mixed-effects model for each class in each training fold. With *K* classes and *F* cross-validation folds, each model specification requires *K* × *F* mixed-model fits. Prediction is cheaper because each test subject is scored by evaluating a multivariate normal density whose dimension is the number of observed visits for that subject. In sparse data, this dimension is small. The method is therefore practical for the study sizes considered here. The present analysis reports pooled held-out cross-validation metrics, not formal confidence intervals. For confirmatory clinical studies, repeated cross-validation, bootstrap intervals, or an external test cohort would be needed to quantify uncertainty more fully. The quadratic specification is a compromise between flexibility and stability. Linear models may miss curved progression patterns, while cubic models, splines, or Gaussian processes may be unstable when class-specific sample sizes are small. We use AIC and BIC diagnostics in the Parkinson’s dense histories to check whether a quadratic model is a reasonable parsimonious choice [2, 35]. The number of trajectory phenotypes is also supported by clustering diagnostics, including inertia and silhouette values [22, 31]. These diagnostics are not presented as proof of a unique biological grouping; they are used to make the sparse recovery experiment transparent and reproducible.

### 3.8 Experimental setup

The Parkinson’s application in this paper is framed carefully. The outcome labels are not independent clinical diagnoses, physician-assigned stages, or externally validated progression endpoints. Instead, dense motor-UPDRS histories are used to define data-derived mathematical trajectory phenotypes. Most observations are then hidden, and the classifier is asked to recover the phenotype from the remaining sparse visits. This makes the application an internal sparse trajectory recovery experiment. It is useful as an algorithmic stress test because it asks whether the method can recover a known trajectory structure under severe sparsification. It should not be interpreted as proof of clinical diagnostic validity. That distinction is important because digital biomarker research in PD is moving quickly, including work on speech, gait, smartphone and smartwatch data, wearable sensors, computer vision, and deep learning models for progression and severity monitoring [23, 32, 34]. Clinical claims require independent outcomes, external cohorts, and prospective validation. Evaluation focuses on accuracy, macro-F1, Matthews correlation coefficient (MCC), and macro-AUC because class imbalance can make accuracy alone misleading [11, 24, 27, 38].

## 4 Results

The results section is organized into two parts: the simulation study and the Parkinson’s disease sparse trajectory recovery analysis. All model-comparison tables report held-out subject-level results from stratified 5-fold cross-validation unless otherwise noted.

### 4.1 Simulation study

The simulation study was designed to evaluate the proposed classifier in a controlled setting where the true trajectory classes are known using stratified subject-level 5-fold cross-validation [10, 17]. We generated sparse longitudinal data sets with *N* = 50, 100, 300, 500, and 1000 subjects. For each sample size, subjects were assigned to three classes using an imbalanced 1:2:3 ratio, matching a common biomedical setting in which one trajectory group is more common than the others [27, 38]. For example, the *N* = 300 setting contains 50, 100, and 150 subjects in Classes 0, 1, and 2, respectively, while the other sample sizes preserve the same imbalance as closely as possible using integer class counts.

Each simulated subject has between 4 and 8 visits over a two-year interval, and visit times are sampled independently from a continuous uniform distribution [19, 41]. The class-specific mean curves are quadratic and include crossing or curved patterns, so simple averages and linear slopes are not sufficient to fully distinguish the groups. Subject-specific random intercept variation and independent Gaussian measurement noise are added to mimic between-subject heterogeneity and measurement error [10, 41]. The simulation is not intended to reproduce every feature of a clinical PD study. Instead, it isolates the statistical challenge that motivates the proposed method: sparse, irregular, multiclass longitudinal classification when class membership is encoded in trajectory shape.

Table 1 summarizes the simulation sample-size design. Table 2 reports the held-out performance of the proposed multiclass MLC model across the five sample sizes. The same subject-level fivefold cross-validation protocol is used for each setting, and all metrics are computed from held-out subjects rather than from individual visits [17, 25].

**Table 1:** Simulation design for the sample-size experiment. Class sizes preserve the original 1:2:3 imbalance.

| Subjects | Class 0 | Class 1 | Class 2 | Mean observations | Mean obs/subject |
| --- | --- | --- | --- | --- | --- |
| 50 | 8 | 17 | 25 | 305.0 | 6.10 |
| 100 | 17 | 33 | 50 | 597.0 | 5.97 |
| 300 | 50 | 100 | 150 | 1815.0 | 6.05 |
| 500 | 83 | 167 | 250 | 2991.0 | 5.98 |
| 1000 | 167 | 333 | 500 | 5988.0 | 5.99 |

**Table 2:**
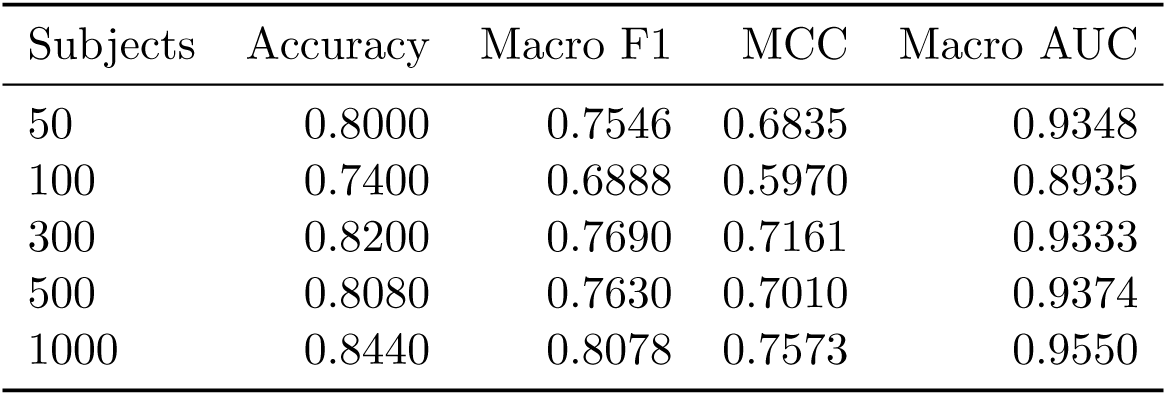
Proposed multiclass MLC performance across simulated sample sizes using held-out subject-level stratified 5-fold cross-validation.

| Subjects | Accuracy | Macro F1 | MCC | Macro AUC |
| --- | --- | --- | --- | --- |
| 50 | 0.8000 | 0.7546 | 0.6835 | 0.9348 |
| 100 | 0.7400 | 0.6888 | 0.5970 | 0.8935 |
| 300 | 0.8200 | 0.7690 | 0.7161 | 0.9333 |
| 500 | 0.8080 | 0.7630 | 0.7010 | 0.9374 |
| 1000 | 0.8440 | 0.8078 | 0.7573 | 0.9550 |

The sample-size results show that the proposed model remains competitive across all simulated cohort sizes, with macro-AUC consistently above 0.89. Performance is not perfectly monotone because each row is generated under a sparse irregular sampling design with random subject-level effects and fixed five-fold partitions. The main pattern is that larger samples stabilize the likelihood-based classifier, with the *N* = 1000 setting achieving accuracy 0.8440, macro-F1 0.8078, MCC 0.7573, and macro-AUC 0.9550.

A full model-comparison study was also conducted at *N* = 300 to compare the proposed method with statistical and machine-learning baselines under the same simulated trajectory setting. Table 3 reports this comparison. The proposed multiclass MLC model achieved the best overall performance, with accuracy 0.8400, macro-F1 0.8099, MCC 0.7570, and macro-AUC 0.9575. The linear MLC baseline was weaker, suggesting that the curvature term and random slope helped recover the simulated class structure. Functional kNN performed reasonably well and was stronger than the scalar-summary random forest and SVM on several metrics, but it did not match the proposed likelihood model on macro-F1 or MCC.

**Table 3:** Held-out subject-level stratified 5-fold cross-validation metrics for Synthetic Simulation Data (N=300 full model comparison).

| Model | Accuracy | Macro-F1 | MCC | Macro-AUC |
| --- | --- | --- | --- | --- |
| Baseline MLC (linear) | 0.6800 | 0.6560 | 0.5231 | 0.8563 |
| Multiclass MLC (quadratic) | 0.8400 | 0.8099 | 0.7570 | 0.9575 |
| Functional kNN | 0.7733 | 0.6696 | 0.6204 | 0.8697 |
| Random forest | 0.6467 | 0.5405 | 0.4144 | 0.8034 |
| Support vector machine | 0.6300 | 0.6136 | 0.4656 | 0.8402 |

The simulation results support the main methodological claim: when class information is carried by sparse nonlinear trajectories, a likelihood model that directly represents trajectory shape can outperform models based on coarse summaries [14, 17, 42]. The results also show why reporting only accuracy is not sufficient. Some models may classify the larger class reasonably well while doing less well on smaller classes. Macro-F1 and MCC make this imbalance visible [24, 38].

### 4.2 Case study on Parkinson’s disease prediction

The real-data application uses the Parkinson’s telemonitoring motor-UPDRS data, a public longitudinal data set of repeated clinical motor scores and related measurements collected for remote monitoring research [40]. The analysis includes *N* = 42 subjects and 5,875 dense observations. Dense histories are first used to define three trajectory phenotypes by fitting subject-level polynomial descriptors and applying *k*-means clustering. The resulting phenotype class sizes are 10, 10, and 22 subjects. Sparse recovery inputs are then generated by retaining only a subset of each subject’s observations, leaving 587 observations and hiding approximately 90% of the original dense measurements. The real-data task should be interpreted as an internal sparse trajectory recovery experiment [40]. The target labels are data-derived mathematical phenotypes rather than independently assigned clinical outcomes. Therefore, the analysis tests whether a classifier can recover a dense-history trajectory group from sparse observations of the same longitudinal process. It does not test whether the method diagnoses PD, predicts clinical stage, or generalizes to externally assigned progression labels. This distinction is important because motor-UPDRS is a clinical rating scale with bounded and ordinal features, whereas the main likelihood model uses a Gaussian continuous approximation. The Gaussian approximation is useful for developing and testing the multiclass likelihood contrast framework, but it is not the final form that should be used for confirmatory clinical modeling [1, 8]. Table 4 reports the Parkinson’s sparse recovery results. The proposed multiclass MLC model achieved accuracy 0.8095, macro-F1 0.8160, MCC 0.7107, and macro-AUC 0.9268. Functional kNN achieved the highest macro-AUC, indicating strong ranking performance, but the likelihood model gave better final class assignments by evaluation matrices. The random forest and support vector machine baselines were weaker, which is consistent with the fact that their input summaries do not fully represent curved longitudinal histories.

**Table 4:** Held-out subject-level stratified 5-fold cross-validation metrics for Parkinson’s Sparse Trajectory Recovery.

| Model | Accuracy | Macro-F1 | MCC | Macro-AUC |
| --- | --- | --- | --- | --- |
| Baseline MLC (linear) | 0.5238 | 0.4470 | 0.2241 | 0.7087 |
| Multiclass MLC (quadratic) | 0.8095 | 0.8160 | 0.7107 | 0.9268 |
| Functional kNN | 0.7857 | 0.7490 | 0.6635 | 0.9417 |
| Random forest | 0.6429 | 0.5145 | 0.3908 | 0.7395 |
| Support vector machine | 0.6429 | 0.5724 | 0.4182 | 0.7499 |

Table 5 provides an additional training, validation, and held-out test diagnostic for the proposed Parkinson’s sparse recovery model. The diagnostic uses the same subject-level splitting principle, with a validation subset drawn only from the training portion of each fold. These values are not used to tune the final model; they are included to show that the validation and held-out test results are close to the training-fold performance and do not suggest severe overfitting.

**Table 5:** Training, validation, and held-out test performance for the proposed multiclass MLC model in the Parkinson’s sparse recovery experiment. The diagnostic uses stratified subject-level splitting with an approximate 64%/16%/20% train/validation/test structure within the 5-fold workflow.

| Model | Accuracy | Macro-F1 | MCC | Macro-AUC |
| --- | --- | --- | --- | --- |
| Training | 0.8690 | 0.8723 | 0.7962 | 0.9517 |
| Validation | 0.8000 | 0.7980 | 0.7276 | 0.9441 |
| Test | 0.8095 | 0.8160 | 0.7107 | 0.9268 |

Figure 2 compares the predictive performance of the proposed multiclass MLC model across the controlled simulation environment and the Parkinson’s sparse trajectory recovery experiment. The plot is intentionally compact, focusing exclusively on the proposed model to illustrate its algorithmic robustness when transitioning from synthetic data to a heavily sparsified empirical dataset. Crucially, the method demonstrates stability in macro-F1 and MCC across both settings, which are critical metrics when evaluating datasets with unequal class sizes. This consistency highlights the model’s capability for algorithmic trajectory recovery from limited, unaligned observations, successfully stress-testing the framework prior to future prospective clinical validation.

**Figure 2:**
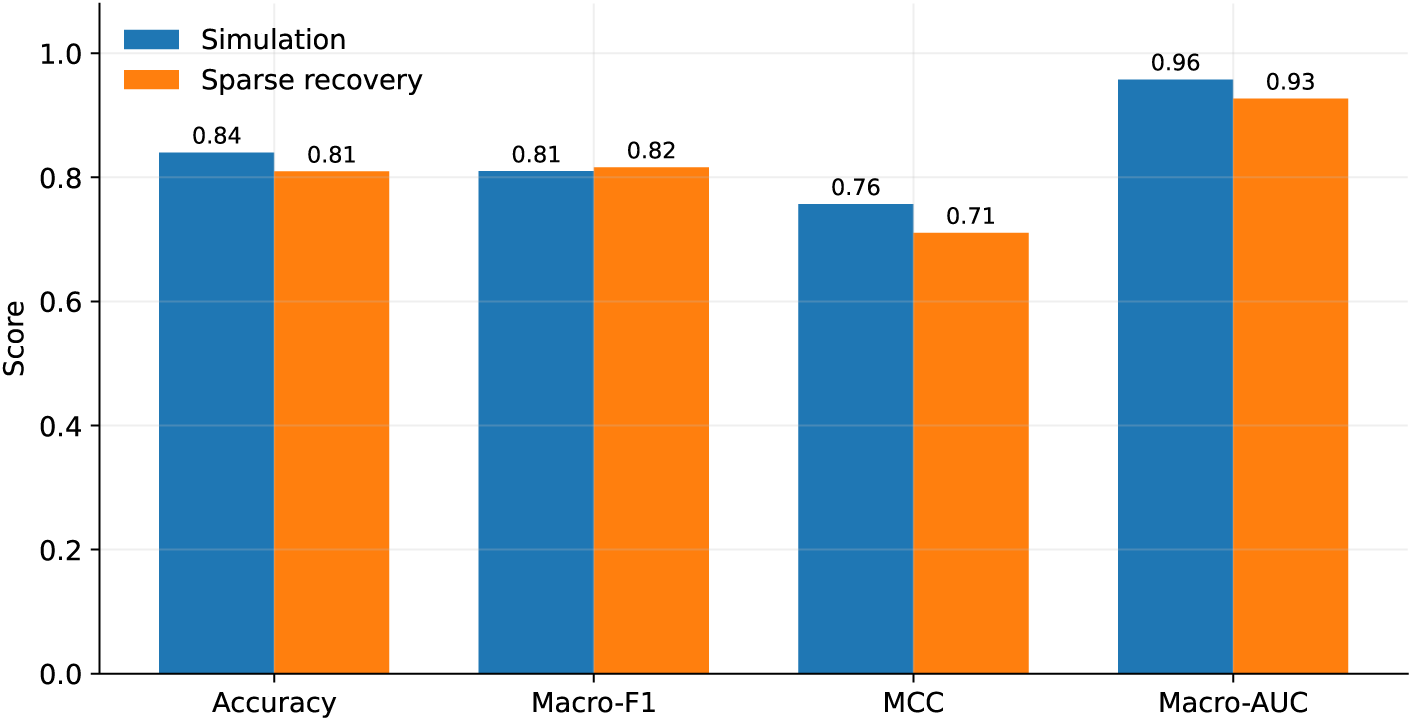
Multiclass MLC performance across the simulation and Parkinson’s sparse trajectory recovery experiments. The figure reports accuracy, macro-F1, MCC, and macro-AUC for the proposed quadratic mixed-likelihood classifier.

Design diagnostics were used to check the Parkinson’s sparse recovery setup before classification. The clustering summaries supported a practical three-phenotype solution, although they do not prove that three is the only possible number of progression groups. Dense-history model-selection diagnostics also showed the expected AIC-BIC tradeoff: the spline specification was more flexible, while the quadratic model was preferred by BIC as the more parsimonious mean trajectory model [2, 35].These diagnostics support the use of a quadratic main analysis and suggest that spline-based likelihood contrasts may be worth exploring in larger cohorts, but they are treated as design checks rather than as evidence for a unique biological taxonomy

## 5 Discussion

In this study, we developed and evaluated a multiclass likelihood contrast learning framework for classification of sparse and irregularly observed longitudinal trajectories. Across both simulation experiments and a Parkinson’s disease sparse trajectory recovery application, the proposed approach demonstrated strong and consistent performance, achieving high accuracy, macro-F1, MCC, and macro-AUC while maintaining an interpretable statistical structure[10, 17]. These findings suggest that likelihood-based artificial intelligence methods can effectively recover clinically relevant longitudinal patterns without requiring observations to be aligned or transformed into fixed-length feature vectors[19, 41].

A key finding of this study is that preserving trajectory shape substantially improves classification performance. In the simulation study, the proposed model outperformed random forest, support vector machine, and linear mixed-likelihood baselines, particularly in macro-F1 and MCC, indicating improved recovery of minority trajectory classes [6, 9, 24]. Similar findings were observed in the Parkinson’s disease application, where the proposed framework achieved a macro-F1 score of 0.8160 and MCC of 0.7107 despite approximately 90% of observations being removed. These results suggest that sparse longitudinal records continue to contain meaningful information about disease progression when analyzed using models that explicitly account for temporal structure[29, 42].

The findings also highlight the potential role of interpretable AI in longitudinal clinical research. Recent advances in artificial intelligence for Parkinson’s disease have focused on deep learning approaches applied to speech, gait, wearable sensors, and electronic health records. Although these methods can achieve strong predictive performance, they often require large datasets and may provide limited insight into how predictions are generated[34, 37]. In contrast, the proposed framework is based on class-specific mixed-effects trajectories and likelihood comparisons, allowing predictions to be directly linked to clinically interpretable progression patterns[3, 17]. This transparency may be particularly valuable in biomedical settings where understanding disease trajectories is as important as prediction accuracy.

An additional strength of the proposed approach is its ability to accommodate irregular followup schedules. Longitudinal studies of Parkinson’s disease and other chronic conditions frequently contain unequal numbers of observations, missed visits, and subject-specific assessment times[10, 19]. Many conventional machine-learning methods require substantial preprocessing, interpolation, or feature engineering before such data can be analyzed. By directly modeling repeated measurements through mixed-effects likelihoods, the proposed framework naturally handles irregular observation schedules while preserving within-subject temporal information[41].

Several limitations should be considered when interpreting these findings. First, the Parkinson’s disease application represents a sparse trajectory recovery experiment rather than a clinical prediction study. The target labels were derived from data-driven trajectory phenotypes constructed from dense motor-UPDRS histories and do not represent externally validated clinical outcomes[26, 40]. Consequently, the reported performance metrics should be interpreted as evidence of successful recovery of latent trajectory structure rather than diagnostic or prognostic accuracy. Second, the study included only 42 subjects, resulting in limited sample sizes for some trajectory classes. Although subject-level cross-validation and train-validation-test diagnostics suggested stable performance, larger external cohorts are needed to evaluate generalizability[27, 38]. Third, the current implementation relies on Gaussian mixed-effects models, whereas motor-UPDRS scores are ordinal and bounded. Future studies should investigate ordinal generalized linear mixed-model likelihoods that more closely reflect the underlying measurement scale. Finally, the present work focused on a single longitudinal outcome, whereas contemporary Parkinson’s disease studies increasingly incorporate multimodal biomarkers, including speech features, wearable sensors, gait measurements, imaging data, and digital health assessments[23, 32, 34].

These findings have broader implications beyond Parkinson’s disease. Many clinical conditions, including Alzheimer’s disease, multiple sclerosis, cancer progression, and cardiovascular disease, are characterized by sparse and irregular longitudinal follow-up[39]. The proposed likelihood contrast framework provides a general strategy for leveraging longitudinal information while maintaining interpretability and computational efficiency. As healthcare datasets continue to expand in temporal depth, methods that combine the transparency of statistical modeling with the predictive capabilities of artificial intelligence may become increasingly important for clinical decision support and patient stratification[39].

Overall, this study demonstrates that multiclass likelihood contrast learning can effectively recover sparse longitudinal trajectory patterns and offers a competitive, interpretable alternative to more complex machine-learning approaches. Although further validation using externally assigned clinical outcomes and larger prospective cohorts is required, the results suggest that likelihood-based AI may provide a practical framework for modeling disease progression in Parkinson’s disease and other chronic disorders characterized by irregular longitudinal data[40, 41].

Although our proposed framework works well for both simulation study and prediction of Parkinson’s disease progression, several limitations should be considered[26, 40]. First, the Parkinson’s disease application was designed as a sparse trajectory recovery study in which progression phenotypes were derived from dense longitudinal histories. Accordingly, the reported performance reflects recovery of longitudinal progression patterns rather than clinical diagnosis or prognosis. Future studies should evaluate the framework using externally assigned clinical outcomes and independent cohorts. Second, the current implementation uses Gaussian mixed-effects models for motor-UPDRS trajectories. Although this approximation performed well in practice, future work could extend the likelihood contrast framework to ordinal generalized linear mixed models that more closely align with the ordinal nature of clinical rating scales. Third, the Parkinson’s analysis involved a relatively small cohort, and larger multicenter studies will be important for assessing generalizability across patient populations, follow-up schedules, and clinical settings. Finally, this study focused on a single longitudinal outcome. The proposed framework is naturally extensible to multimodal longitudinal data, including speech, gait, wearable-sensor, and digital biomarker measurements, which may further improve characterization of Parkinson’s disease progression[23, 32, 34].

### Public Health and Clinical Implications

As longitudinal clinical and digital health data become increasingly available, methods capable of analyzing sparse and irregular follow-up records will be essential for precision medicine and population health management [39]. The proposed likelihood contrast framework offers a practical and interpretable approach for identifying disease progression patterns from incomplete longitudinal data, a common challenge in real-world clinical practice. In Parkinson’s disease, such models could support earlier recognition of progression subtypes, facilitate personalized monitoring schedules, and improve patient selection for clinical trials and targeted interventions. More broadly, the framework may be applicable to other chronic diseases characterized by heterogeneous progression and irregular follow-up, including Alzheimer’s disease, multiple sclerosis, cardiovascular disease, and cancer survivorship. By leveraging routinely collected longitudinal data without requiring complex preprocessing or highly parameterized deep-learning architectures, this approach has the potential to enhance clinical decision support while remaining transparent and accessible to healthcare practitioners.

## 6 Conclusion

This study presents a multiclass likelihood contrast learning framework for classification of sparse and irregularly observed longitudinal trajectories. In both simulation studies and a Parkinson’s disease motor-UPDRS application, the proposed approach demonstrated strong performance while preserving the temporal structure of repeated measurements through class-specific mixed-effects models. Notably, the framework successfully recovered trajectory phenotypes even after approximately 90% of longitudinal observations were removed, highlighting its ability to extract meaningful progression patterns from limited follow-up data. Beyond predictive performance, the proposed method offers an interpretable alternative to more complex artificial intelligence approaches. By directly modeling longitudinal disease trajectories and assigning class membership through likelihood contrasts, the framework provides clinically understandable predictions while naturally accommodating irregular visit schedules and unequal numbers of observations per patient. Although the Parkinson’s disease analysis focused on recovery of data-derived progression phenotypes rather than externally validated clinical outcomes, the findings suggest that likelihood-based AI may serve as a practical tool for longitudinal patient stratification and disease monitoring. In clinical settings, such models could support identification of progression subgroups, facilitate risk-based follow-up scheduling, and assist in selecting patients for targeted interventions or clinical trials. Future research should evaluate the framework in larger multicenter cohorts using independently assigned clinical outcomes, prospectively collected data, and multimodal longitudinal biomarkers, including speech, gait, wearable-sensor, and digital health measures. Extensions to ordinal generalized linear mixed models may further improve alignment with clinical rating scales such as the MDS-UPDRS [12, 26]. As longitudinal clinical data become increasingly available through remote monitoring and digital health platforms, interpretable AI approaches such as likelihood contrast learning may provide an important bridge between advanced predictive modeling and clinically actionable decision support [34, 40].

## Data Availability

All data produced in the present work are contained in the manuscript

## Acknowledgment

The authors thank the researchers and data contributors whose publicly available Parkinson’s disease longitudinal data made this study possible. Authors are also thankful to Grammarly for writing support.

## Data and code availability

The Parkinson’s data analyzed in this paper are publicly available from the Parkinson’s telemonitoring study originally described by Tsanas et al. [40]. Code will be available upon request.

## Ethics statement

This study uses simulated data and a publicly available de-identified Parkinson’s telemonitoring data set. No new human-subject data were collected for this methodological analysis. Any future clinical deployment or prospective validation of the proposed classifier would require appropriate institutional review, data-governance review, and evaluation of fairness, calibration, and safety in the intended use population.

## Funding

No external funding was received for this study.

## Competing Interest

The authors declare no competing interests.

## Notes

### Competing Interest Statement

The authors have declared no competing interest.

